# Neutralising antibody activity against SARS-CoV-2 variants, including Omicron, in an elderly cohort vaccinated with BNT162b2

**DOI:** 10.1101/2021.12.23.21268293

**Authors:** Joseph Newman, Nazia Thakur, Thomas P. Peacock, Dagmara Bialy, Ahmed ME Elreafey, Carlijn Bogaardt, Daniel L. Horton, Sammy Ho, Thivya Kankeyan, Christine Carr, Katja Hoschler, Wendy S. Barclay, Gayatri Amirthalingam, Kevin Brown, Bryan Charleston, Dalan Bailey

**Affiliations:** The Pirbright Institute, Guildford, Surrey, GU24 0NF, United Kingdom; Nuffield Department of Medicine, The Jenner Institute, Oxford, OX3 7DQ, United Kingdom; Department of Infectious Disease, Imperial College – London, W2 1PG, United Kingdom; UK Health Security Agency (UKHSA), UK; Department of Pathology and Infectious Diseases, School of Veterinary Medicine, University of Surrey

## Abstract

SARS-CoV-2 variants threaten the effectiveness of tools we have developed to mitigate against serious COVID-19. This is especially true in clinically vulnerable sections of society including the elderly. Using sera from BNT162b2 (Pfizer–BioNTech) vaccinated individuals aged between 70 and 89 (vaccinated with two doses 3-weeks apart) we examined the neutralising antibody (nAb) response to wildtype SARS-CoV-2. Between 3 and 20-weeks post 2^nd^ dose, nAb titres dropped 4.9-fold to a median titre of 21.3 (ND80) with 21.6% of individuals having no detectable nAbs at the later time point. Experiments examining the neutralisation of twenty-one different SARS-CoV-2 variant spike proteins confirmed a significant potential for antigenic escape, especially for the Omicron (BA.1), Beta (B.1.351), Delta (B.1.617.2), Theta (P.3), C.1.2 and B.1.638 variants. Interestingly, however, the recently-emerged sub-lineage AY.4.2 was more efficiently neutralised than parental Delta pseudotypes. Combining pseudotype neutralisation with specific receptor binding domain (RBD) ELISAs we confirmed that changes to position 484 in the spike RBD were predominantly responsible for SARS-CoV-2 nAb escape, although the effect of spike mutations is both combinatorial and additive. Lastly, using sera from the same individuals boosted with a 3^rd^ dose of BNT162b2 we showed that high overall levels of neutralising antibody titre can provide significant levels of cross-protection against Omicron. These data provide evidence that SARS-CoV-2 neutralising antibodies wane over time and that antigenically variable SARS-CoV-2 variants are circulating, highlighting the importance of ongoing surveillance and booster programmes. Furthermore, they provide important data to inform risk assessment of new SARS-CoV-2 variants, such as Omicron, as they emerge.

## Introduction

The effects of the COVID-19 pandemic have, in some countries, been mitigated by the implementation of highly efficacious vaccines, which have reduced hospitalisations and deaths. In late 2020 and early 2021 this was widely achieved (in the UK, France and elsewhere) through vaccination with BNT162b2 (Pfizer–BioNTech) *–* a lipid nanoparticle-formulated, nucleoside-modified RNA vaccine encoding prefusion stabilized SARS-CoV-2 spike; or ChAdOx1 nCoV-19 (AZD1222, Oxford–AstraZeneca), an adenoviral-vectored vaccine expressing wild-type (non-stabilised) spike [1, 2]. Initially in the UK, for BNT162b2, two doses of this vaccine were administered 3-weeks apart, with many of the most clinically vulnerable (within the nine priority groups established by the UK’s Joint Committee on Vaccination and Immunisation [JCVI]) receiving their vaccines with this dosing interval. However, in the UK, this schedule was quickly changed to ‘up to 12-weeks’, to maximise use of limited supplies of these vaccines and to protect the largest possible number of people from developing serious disease. This remained the strategy as vaccination was extended to the priority groups further down JCVI’s list (stratification based primarily on age), before being opened up to all adults later in 2021, as well as children over 12. Third doses, as well as boosters, are also now available to all adults [3]. To date, detailed information on vaccine responses in elderly populations (>65) vaccinated with BNT162b2 3-weeks apart is lacking, in particular data on neutralising antibody (nAb) titres over time, correlations between ELISA and nAb titres, cross-protective nAb titres against SARS-CoV-2 variants, and the role of boosters in enhancing this cross protection. These data are relevant to elderly cohorts in the UK, but also internationally where the 3-week interval between doses is followed. Neutralising antibodies in the elderly are of especial significance, as it is well established that vaccine responses in this demographic are less robust [4, 5], and this group may represent a vulnerable cohort for SARS-CoV-2 variants.

Whereas the SARS-CoV-2 virus that caused the first global wave had little genetic, antigenic or other phenotypic diversity, subsequent waves comprised extremely diverse SARS-CoV-2 ‘variants’, defined by genetic, antigenic and phenotypic divergence from preceding strains [6]. The most widespread or concerning of these were classified by the World Health Organisation (WHO) and/or UKHSA as ‘variants of concern (VOC)’, ‘variants of interest/variants under investigation’ (VOI, WHO; VUI, UKHSA) or ‘variants under monitoring’ VUM; these VOCs and VUIs were subsequently given Greek letter identifiers [7]. The first described of these, the Alpha variant (Pango lineage B.1.1.7), emerged in the UK around Autumn 2020 and showed higher transmissibility than previous variants [8, 9]. The VOC Beta/B.1.351 was detected at a similar time in South Africa, with the VOC Gamma/P.1 identified in Brazil not long after. All three of these VOCs showed antigenic distance from the original strain, the spike protein of which is used as the immunogen in BNT162b2 and ChAdOx1 nCoV-19 vaccines [10, 11]. From the start of 2021 many further variants, arose throughout the world, including B.1.1.318, C.36.3, P.3, Mu/B.1.621, B.1.620, B.1.617.3, Lambda/C.37, A.30, AT.1, B.1.638, and C.1.2, with some being designated as VUIs/VUMs, [12-14]. In April 2021 a new variant, Delta (B.1.617.2), rapidly spread across the world after its initial detection in India [15, 16]. Although Delta shows only modest antigenic divergence from the original SARS-CoV-2 strains it has the highest transmissibility of any variant studied to date. Indeed, by November 2021, Delta had largely displaced all other variants and comprises nearly all global sequences, with small, localised pockets of Alpha, Gamma, Mu, B.1.1.318 and C.1.2 remaining but slowly declining [14]. However, a Delta sub-lineage, AY.4.2, has recently arisen in the UK and shows tentative signs of increased prevalence, gradually replacing other Delta sub-lineages [13]. In late November 2021 the Omicron (BA.1, initially denoted B.1.1.529) variant was first detected in southern Africa (specifically Botswana and South Africa), with isolations found across the globe in the following days and weeks. Early data on increased transmissibility, community displacement of Delta and evidence for significant antigenic variation support its immediate classification as a VOC (Omicron) [17-20].

To examine antibody levels and T-cell responses following the extension to the COVID-19 vaccine schedule the UKHSA (formerly Public Health England, PHE) initiated a prospective longitudinal audit of vaccinated adults (the CONSENSUS study). Within CONSENSUS, a cohort of volunteers aged between 70 and 89 received the BNT162b2 vaccine 3-weeks apart. In our study, using sera from 37 individuals in this cohort, we investigated the impact of waning immunity on neutralisation of SARS-CoV-2, the correlation between ELISA, RBD-ELISA and nAb titres, as well as the impact of various SARS-CoV-2 VOCs and VUMs/VUIs on viral neutralisation. These data indicate that this clinically vulnerable priority group may be especially susceptible to repeat infection with SARS-CoV-2 variants that are antigenically distinct from the originally emerged strain; however, a 3^rd^ dose of BNT162b2 can mitigate against this risk. This susceptibility is the result of low overall nAb titres and appears to be mechanistically defined by specific changes to the SARS-CoV-2 spike RBD, in particular E484 changes. These data provide a key tool for risk assessing current and future SARS-CoV-2 variants, including Omicron/BA.1 or sub-lineages (BA.2).

## Results

Using a SARS-CoV-2 spike pseudotype-based micro-virus neutralisation assay (mVNT), we determined neutralisation titres (ND80s) in 37 UK-based participants (median age 78 years [IQR 75-80]) who had been vaccinated with BNT162b2 (Pfizer–BioNTech) 3-weeks apart (median 21 days [IQR 21-21]). Titres from samples taken at 3 (n=37, median 22 days [IQR 22-23] and 20-weeks (n=35, median 135 days [IQR 134-136.5]) post 2^nd^ dose immunisations were initially evaluated with D614-based (wild-type [WT]/Wuhan) pseudotypes to match the BNT162b2 immunogen. Dividing the cohort by age into individuals aged 70-79 (n=24, median age 77 years [IQR 73.5-80]) and 80-89 years old (n=13, median age 81 years [IQR 80-84]) we observed median titres (ND80) of ≤128.1 (IQR 28.03-231.2) and ≤62.6 (IQR 25.45-129.6), respectively, at 3-weeks post-vaccination. At 20-weeks post-vaccination these titres had dropped to ≤24.84 (ages 70-79; IQR 14.59-61.46) and ≤16.0 (ages 80-89; IQR 10-24.34), a median reduction of ≤5.2-fold and ≤3.9-fold, respectively (Figure 1A-B; exemplar RLU data provided in Supplemental figure 1 A-B). At 3-weeks post 2^nd^ dose 3/24 (12.5%) of 70-79 year olds and 3/13 (23.1%) of 80-89 year olds had no detectable neutralising antibodies (ND80 = ≤10; limit of assay detection), with this increasing to 4/22 (18.2%) and 4/13 (30.8%), respectively, at 20-weeks. The 3-week ND80 titres were also converted to IU/ml, based on comparisons to WHO’s international standard for SARS-CoV-2 serological assays (NIBSC code: 20/136) (Supplemental Figure 1C; median titres; ages 70-79, 344.5 IU/ml, 80-89, 168.3 IU/ml). The same sera samples were previously analysed [21] by ELISA (Roche Elecsys anti-SARS-CoV-2 S ECLIA) and there was a strong correlation between mVNT and ELISA titres in both age groups (70-79, Spearman r = 0.84, Figure 1C; 80-89, Spearman r=0.91, Figure 1D).

**Figure 1:**
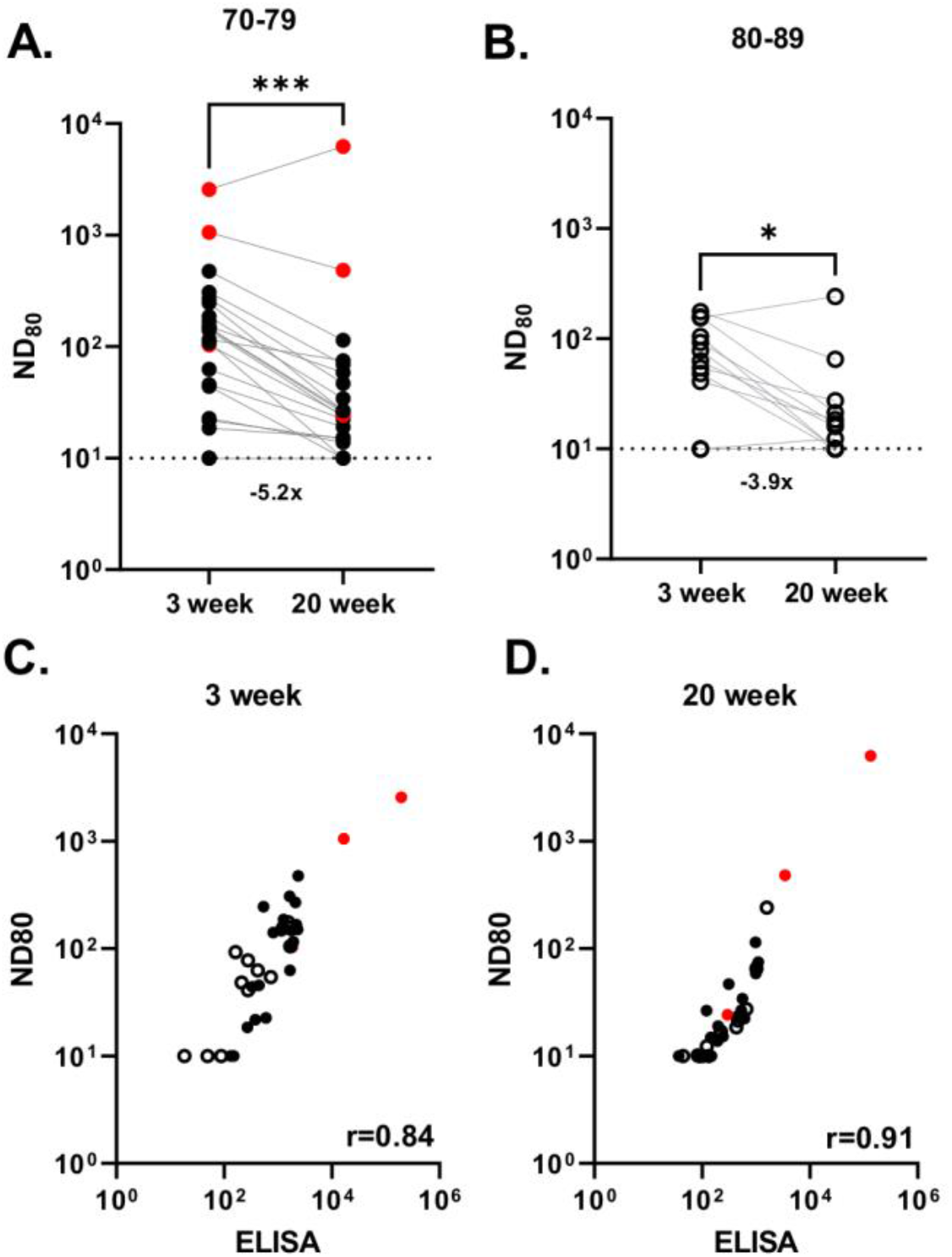
Neutralising antibody responses generated following BNT162b2 vaccination. Neutralisation titres calculated using pseudotypes bearing the SARS-CoV-2 D614 (Wuhan) spike and sera from a cohort of BNT162b2 vaccinated individuals (n=37), recruited as part of the UK CONSENSUS trial, aged 70-79 (n=24, solid circle symbols) **(A)** or 80-89 (n=13, open circle symbols) **(B)**. Symbols in red represents samples taken from individuals who tested positive for SARS-CoV-2 Nucleoprotein by ELISA, indicative of previous infection. Sera was collected from the same individuals at 3- (n=37 total) and 20-weeks (n=35 total) post 2^nd^ dose, with a vaccination interval of 3 weeks between 1^st^ and 2^nd^ doses. Titres are expressed as serum fold-dilution required to achieve 80% virus neutralisation, with the titre (ND80) calculated by *xy* interpolation from the mVNT data series (dilution, *x* versus luciferase activity, RLU, *y*). Statistical comparison of ND80 titres at 3 and 20 weeks was performed using a Wilcoxon matched-pairs signed rank test (*; <0.05; ***, <0.001). Fold changes in median ND80 between 3 and 20 weeks are indicated. The detection limit of the assay is indicated with a dotted line. The correlation between ND80 and S ELISA (Roche S) titres recorded from each volunteer, was examined at 3 (n=37) **(C)** and 20 (n=35) **(D)** weeks post 2^nd^ dose, with statistical analysis of the matrix performed using a nonparametric Spearman correlation (r).

The same 3 weeks post 2^nd^ dose sera were subsequently used to investigate neutralisation of pseudotypes bearing the SARS-CoV-2 spike from VOCs Alpha (B.1.1.7), Beta (B.1.351) and Delta (B.1.617.2), as well as the D614G-containing lineage B.1 (Figure 2A). 4/24 (16.7%) of 70-79 year olds and 5/13 (38.5%) of 80-89 year olds had no detectable neutralising antibodies (ND80 = ≤10; limit of assay detection) to Delta at this time point, while 16/24 (66.7%) and 11/13 (84.6%) of 70-79 and 80-89 year olds, respectively, had no identifiable response to Beta (Figure 2B-C). When compared to D614G (median ND80 ≤333.3; IQR 62.53-642.9), in the 70-79 age group, there was a ≤1.1-fold (median ND80 ≤300.0; IQR 53.89-592.8) reduction in neutralisation of Alpha, a ≤9.0-fold drop with Delta (median ND80 ≤37.0; IQR 12.9-68.6) and a ≤33.3-fold drop with Beta (median ND-80 ≤10.0; IQR 10-16.34) (Figure 2B). In the older age group (ages 80-89) the median titres were D614G ≤139.0 (IQR 39.15-316.2), Alpha ≤136.6 (IQR 43.54-334.3), Delta ≤12.4 (IQR 10-47.31) and Beta ≤10 (IQR 10-10) (Figure 2C). For Delta and Beta this equated to a drop in neutralising titre of 11.2-fold and 13.9-fold, respectively. Neutralising titres to Delta at 20-weeks post-vaccination were slightly lower, consistent with the waning antibody response evidenced in Figure 1 (Supplemental Figure 2A-B). However, there was evidence of affinity maturation in some individuals, especially to the Beta VOC, although the median value remained unchanged (Supplemental Figure 2C-D). VNT assays with replication competent SARS-CoV-2 WT D614 and the Beta VOC also identified a reduction in neutralisation for Beta, with the calculated titres correlating well with the mVNT pseudotype data (Supplemental Figure 3). Comparing the S ELISA (Roche Elecsys anti-SARS-CoV-2 S ECLIA) with VOC ND80 titres we identified a strong correlation for D614G (Spearman r = 0.86), Alpha (r = 0.83) and Delta (r = 0.86) neutralisation titres but a poor correlation with Beta (r = 0.52) (Figure 2D). To investigate the mechanisms underpinning the drop in neutralisation seen for specific VOCs we next performed a targeted ELISA with the same sera and recombinant RBDs reflecting WT SARS-CoV-2, Alpha, Delta and Beta spike sequences. In both the 70-79 and 80-89 age groups there was no significant difference in ELISA titres between WT, Alpha and Delta RBDs (Figure 2E-F); however, there was a significant reduction in binding to the Beta RBD (70-79, 2.9-fold compared to WT; 80-89, 2.2-fold), partially correlating with the mVNT results. The S ELISA and RBD-ELISA data (both based on antigens representing WT SARS-CoV-2 spike sequence) showed a strong correlation (Spearman r = 0.93), indicative of good agreement between the two assays (Figure 2G). However, the correlation between RBD-ELISA and ND80 titres was again poor for the Beta VOC (Spearman r = 0.49), albeit relatively consistent for the WT (r = 0.83), Alpha (r = 0.80) and Delta (r = 0.86) RBD-ELISAs.

**Figure 2:**
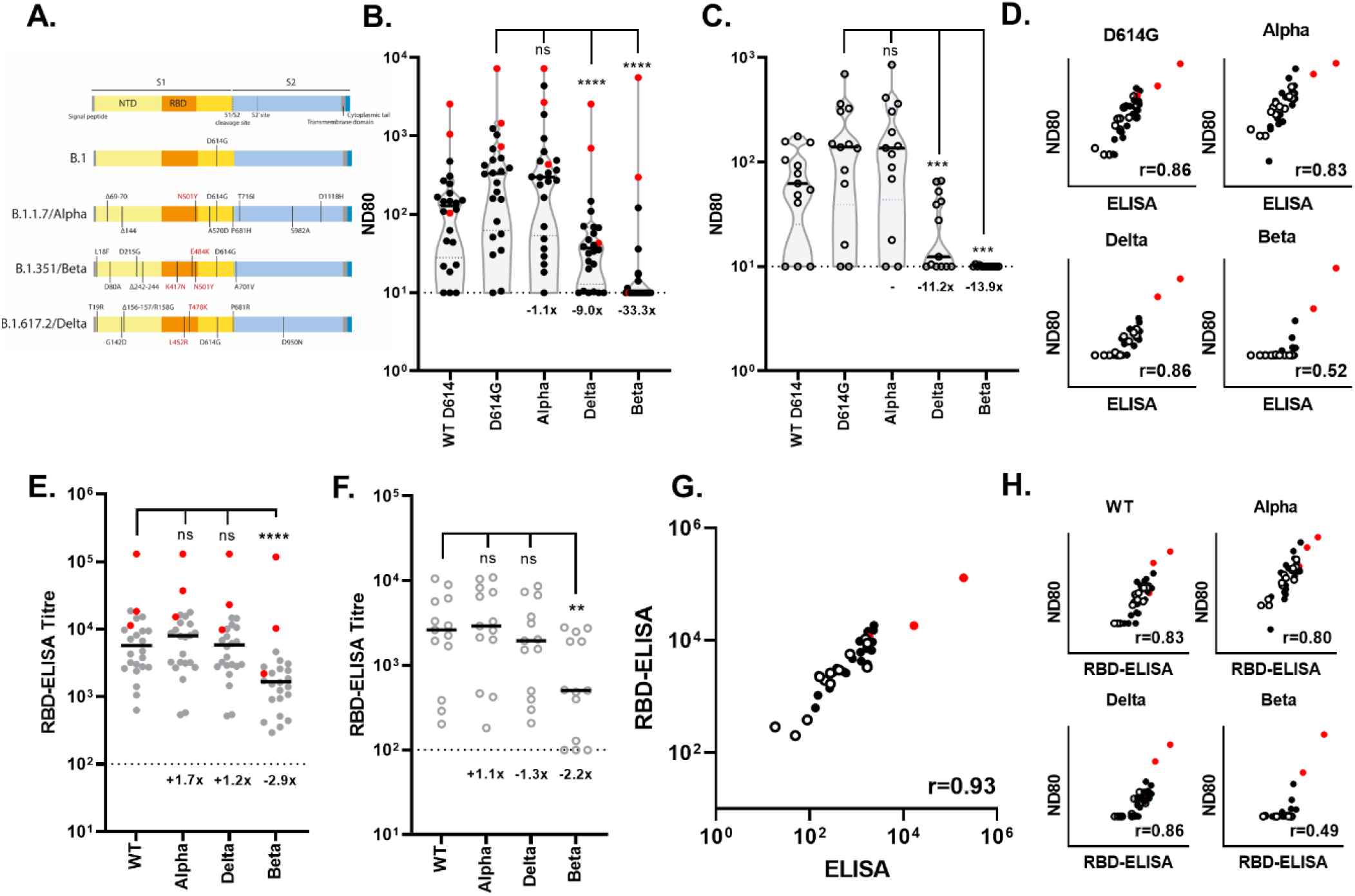
Neutralisation of SARS-CoV-2 variants of concern (VOC) by sera collected from BNT162b2 vaccinated individuals 3 weeks post 2^nd^ dose. **(A)** Schematics illustrating the spike mutation profiles of B.1, B.1.1.7 (Alpha), B.1.617.2 (Delta) and B.1.351 (Beta). Neutralisation of pseudotypes bearing the SARS-CoV-2 WT D614, D614G (B.1), Alpha, Delta or Beta spike were compared in the two age-stratified cohorts, 70-79 (n=24, solid circles) **(B)** and 80-89 (n=13, open circles) **(C)**. Symbols in red represents samples taken from individuals who tested positive for SARS-CoV-2 Nucleoprotein by ELISA, indicative of previous infection. Titres are expressed as serum fold-dilution required to achieve 80% virus neutralisation, with the titre (ND80) calculated by *xy* interpolation from the mVNT data series (dilution, *x* versus luciferase activity, RLU, *y*). Statistical comparison of ND80 titres against the D614G reference was performed using a Wilcoxon matched-pairs signed rank test (***, <0.001; ****, <0.0001). Fold changes in median ND80, compared to D614G are indicated. The detection limit of the assay is indicated with a dotted line. The correlation between ND80 and S ELISA (Roche S) titres recorded for each volunteer (n=37), was then examined **(D)** for each VOC, with statistical analysis of the matrix performed using a nonparametric Spearman correlation (r). The same sera [70-79; n=24, solid circles **(E)** and 80-89; n=13, open circles **(F)**] was analysed with an RBD-based ELISA, using RBDs representing WT (*NB*; as indicated in (A) amino acid 614 is outside the RBD), Alpha, Beta and Delta spikes. Titres are expressed as serum fold-dilution required to achieve a T/N (test OD to negative OD) of 5 (T/N = 5 serves as cut-off for positive samples) by *xy* interpolation from the RBD data series (dilution, *x* versus OD450, *y*). Statistical comparison of RBD-ELISA titres was performed using a nonparametric Friedman test with Dunn’s multiple comparisons of column means (**, <0.005; ****, <0.0001). Fold changes in median titre, compared to WT are indicated. The detection limit of the assay is indicated with a dotted line. The correlation between RBD-ELISA and S ELISA titres (Roche S) **(G)** or ND80 titres for each VOC **(H)** recorded from each volunteer (n=37) was then examined, with statistical analysis of the matrix performed using a nonparametric Spearman correlation (r).

Using a smaller representative pool of sera from the same cohort (3-weeks post 2nd dose; n=16 total; 70-79, n=11; 80-89 n=5), selected by ranking the neutralisation responses of the whole cohort, we widened our analysis to fifteen other SARS-CoV-2 variants, including other VUIs and VUMs, in particular AY.4.2 (Figure 3A). This timepoint was chosen as the titres were higher at 3-rather than 20-weeks post 2^nd^ dose. Several variants showed a significant reduction in neutralisation when compared to D614G (B.1); namely B.1.1.318 (2.9-fold), A.30 (2.0-fold), B.1.617.3 (1.6-fold), B.1.621/Mu (3.0-fold), P.3/Theta (7.2-fold), C.1.2 (10.8-fold), Mu + K417N (3.2-fold), B.1.638 (8.5-fold), AY.4.2 (3.4-fold), and Delta + A222V (2.8-fold) (Figure 3B). Of note, experiments were always performed with a D614G pseudotype control and we showed a good concordance between calculated D614G ND80 titres from individual experiments, highlighting the robust repeatability of our assay and the capacity to compare mVNT titres across data sets (Supplemental Figure 4). Aside from Delta and AY.4.2, which contain the L452R and T478K RBD mutations, all of these variants contain mutations at RBD position 484. Furthermore, for P.3 (Theta), C.1.2 and B.1.638, 4/16 (25.0%), 5/16 and 5/16 (31.3%) of the individuals, respectively, had no detectable neutralising antibodies (ND80 = ≤10; limit of assay detection) to these variants at this time point post-vaccination (for Beta VOC this was 9/16 (56.3%) [calculated from data presented in Figure 2B-C]). Interestingly, the median titres to Delta (≤49.2, IQR 14.24-190.3) were lower than the AY.4.2 (≤82.2, IQR 24.52-82.19) and Delta + A222V (≤100.0, IQR 25.67-211.5) variants, despite the presence of additional mutations in these spike proteins.

**Figure 3:**
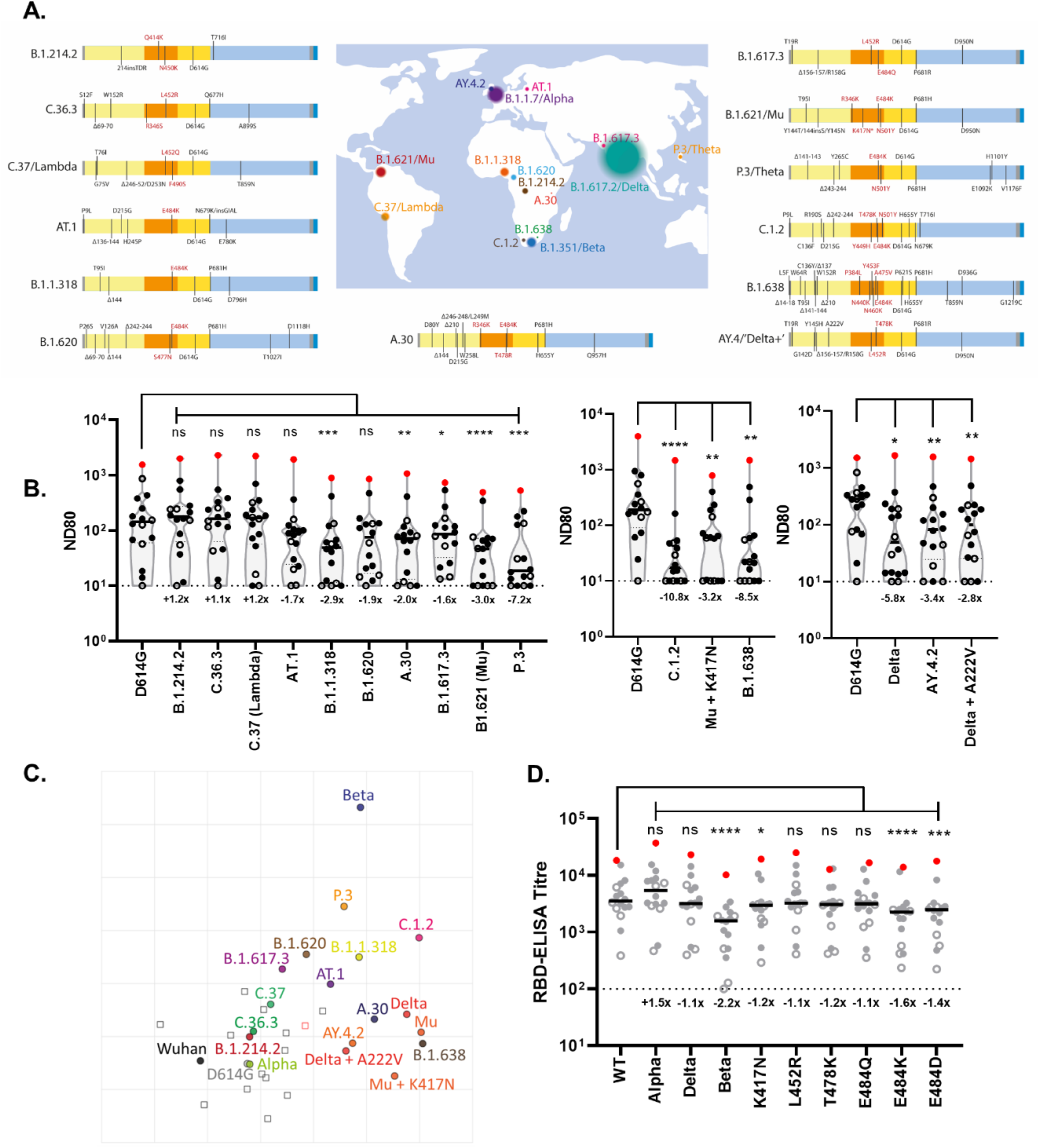
Neutralisation of a broad library of SARS-CoV-2 variants by sera collected from BNT162b2 vaccinated individuals. **(A)** Schematics illustrating the spike mutation profiles of thirteen SARS-CoV-2 variants, variants under investigation (VUIs) or VOCs including B.1, C.37 (Lambda) and B.1.621 (Mu). **(B)** Neutralisation of pseudotypes bearing these spike proteins were compared using a sub-section (n=16) of sera from the BNT162b2-vaccinated (3-week dosing interval, 3-weeks post 2nd dose) age-stratified cohorts (70-79, n=11, solid circles; 80-89 n=5, open circles). The single symbol in red represents a sample taken from an individual who tested positive for SARS-CoV-2 Nucleoprotein by ELISA, indicative of previous infection. Titres are expressed as serum fold-dilution required to achieve 80% virus neutralisation, with the titre (ND80) calculated by *xy* interpolation from the mVNT data series (dilution, *x* versus luciferase activity, RLU, *y*). Statistical comparison of ND80 titres against the D614G reference was performed using a Wilcoxon matched-pairs signed rank test (*, <0.05; **, <0.005, ***, <0.001; ****, <0.0001). The separate graphs represent experiments performed on different days, with each assay including the D614G pseudotype as a reference. Fold changes in median ND80, compared to D614G are indicated. The detection limit of the assay is indicated with a dotted line. **(C)** Two-dimensional antigenic map of variants, based on the ND80 titres in (B). Multidimensional scaling was used to position the sera and variants to best fit target distances derived from the titres. The map is the lowest error solution of 1000 optimisations. Variants are represented by solid circles, sera by open squares (with the Nucleoprotein-ELISA-positive serum in red). Two sera were not used in mapping because of titres consistently below the detection limit. The spacing between grid lines represents one antigenic unit, equivalent to a two-fold dilution in ND80 titres. **(D)** The same sera (70-79, n=11, solid circles; 80-89; n=5, open circles) were analysed with an RBD-based ELISA, using RBDs representing WT, Alpha, Beta and Delta spike (data replotted from Fig.2E/F for comparison) as well as spikes containing the individual mutations K417N, L452R, T478K, E484Q, E484K and E484D (relative to WT). Titres are expressed as serum fold-dilution required to achieve a T/N (test OD to negative OD) of 5 (T/N = 5 serves as cut-off for positive samples) by *xy* interpolation from the RBD data series (dilution, *x* versus OD450, *y*). Statistical comparison of RBD-ELISA titres was performed using a nonparametric Friedman test with Dunn’s multiple comparisons of column means (*, <0.05; ***, <0.001; ****, <0.0001). Fold changes in median titre, compared to WT are indicated. The detection limit of the assay is indicated with a dotted line.

To provide better spatial representation of the antigenic relationships between the different variants, we also performed antigenic cartographic analysis on the collated ND80 titres from a sub-set of the sera (3 weeks post second-dose, n=14) tested against all available VOCs, VUIs and VUMs in mVNTs. Antigenic cartography allows high resolution quantitative comparison and visualisation of antigenic relationships. Antigenic distances between antigens on the map are measured in antigenic units, with one antigenic unit (AU) being the equivalent of a two-fold dilution in titre. The antigenic map of ND80 titres (Figure 3C) again highlights the largest antigenic distance is between D614G and Beta (5.3 AU). Other variants located at an intermediate distance from D614G are C.1.2 (4.0 AU), P.3/Theta (3.5 AU), B.1.621/Mu (3.3 AU), B.1.638 (3.3 AU), Delta (3.1 AU), B.1.1.318 (2.9 AU), Mu + K417N (2.8 AU), A.30 (2.5 AU), B.1.620 (2.3 AU), AT.1 (2.2 AU), AY.4.2 (2.0 AU), B.1.617.3 (1.9 AU), Delta + A222V (1.9 AU), and C.37/Lambda (1.2 AU). The WT/Wuhan (D614) virus, C.36.3, B.1.214.2 and Alpha are all located close (1 AU or less) to D614G. The sera are all located in one part of the map, and not far from Wuhan and D614G viruses; this is as expected from sera from recipients of the same WT/Wuhan spike-based vaccine with similar neutralization profiles. This clustering of sera means interpretation of the distances between the most divergent SARS-CoV-2 variants on the antigenic map (e.g P.3 to Beta) is less reliable than their distances to D614G, demonstrated by the confidence coordination areas for their positions (Supplemental Figure 5A). In addition, to identify the amino acid changes within the RBD responsible for the significant drop in neutralisation in this cohort we repeated the WT, Alpha, Delta and Beta RBD-ELISAs, extending the analysis to include RBDs with single amino acid changes at positions K417, L452, T478 or E484. The only changes that led to a significant drop in binding were K417N (1.2-fold), E484K (1.6-fold) and E484D (1.4-fold), highlighting the importance of these positions in escape from neutralisation (Figure 3D, and antigenic map in Supplemental Figure 5B/C).

In November 2021 the BA.1 (Omicron) VOC was first detected in Southern Africa, rapidly spreading across the globe. By mid-December 2021 infections were highly prevalent in the UK and many other countries. At this time many of the volunteers in the CONSENSUS trial had received their 3^rd^ dose. To understand antigenic changes to the Omicron spike, which has extensive deletions and substitutions (Figure 4A), we performed mVNT assays for all volunteers where a 3^rd^ dose sample was available (n=19; 70-79, n=12; 80-89, n=7). These samples were taken 4 weeks after the booster (median 28 days [IQR 28-28]). At 3 weeks post 2^nd^ dose 10/12 (83%) of 70-79 year olds and 6/7 (86%) of 80-89 year olds had no detectable neutralising antibodies (ND80 = ≤10; limit of assay detection) to Omicron (Figure 4B-C). At 20 weeks post 2^nd^ dose this was 92% and 86%, respectively. However, at 4 weeks following the 3^rd^ dose all volunteers now, significantly, had detectable titres against Omicron (Figure 4B-C). In the 70-79 age group, when compared to D614G (median ND80 ≥530.7; IQR 318.3-883.4), there was a 53.1-fold reduction (median ND80 ≤10; IQR 10-10) in neutralisation of Omicron at 3 weeks post vaccination, but only an 8.0-fold (D614G; median ND80 ≥3070.9; IQR 1498.4-3798.1 vs Omicron; median ND80 384.3; IQR 189.7-735.7) drop 4 weeks after the 3^rd^ dose (Figure 4B). In the older age group (ages 80-89) the median titres for D614G were 1354.1 (IQR 953.3-2103.9) after the 3^rd^ dose, whilst for Omicron this was 80.8 (IQR 59.5-239.2), Figure 4C), a 16.8-fold reduction. Correlating the S ELISA (Roche S1) values for these samples with their Omicron ND80 titres further illustrates the improved neutralisation titres following a 3^rd^ dose of vaccination and also identified an improved correlation between ND80 and ELISA (20 weeks post 2^nd^ dose, Spearman r = 0.53; 4 weeks post 3^rd^ dose, Spearman r=0.82 Figure 4D-E).

**Figure 4:**
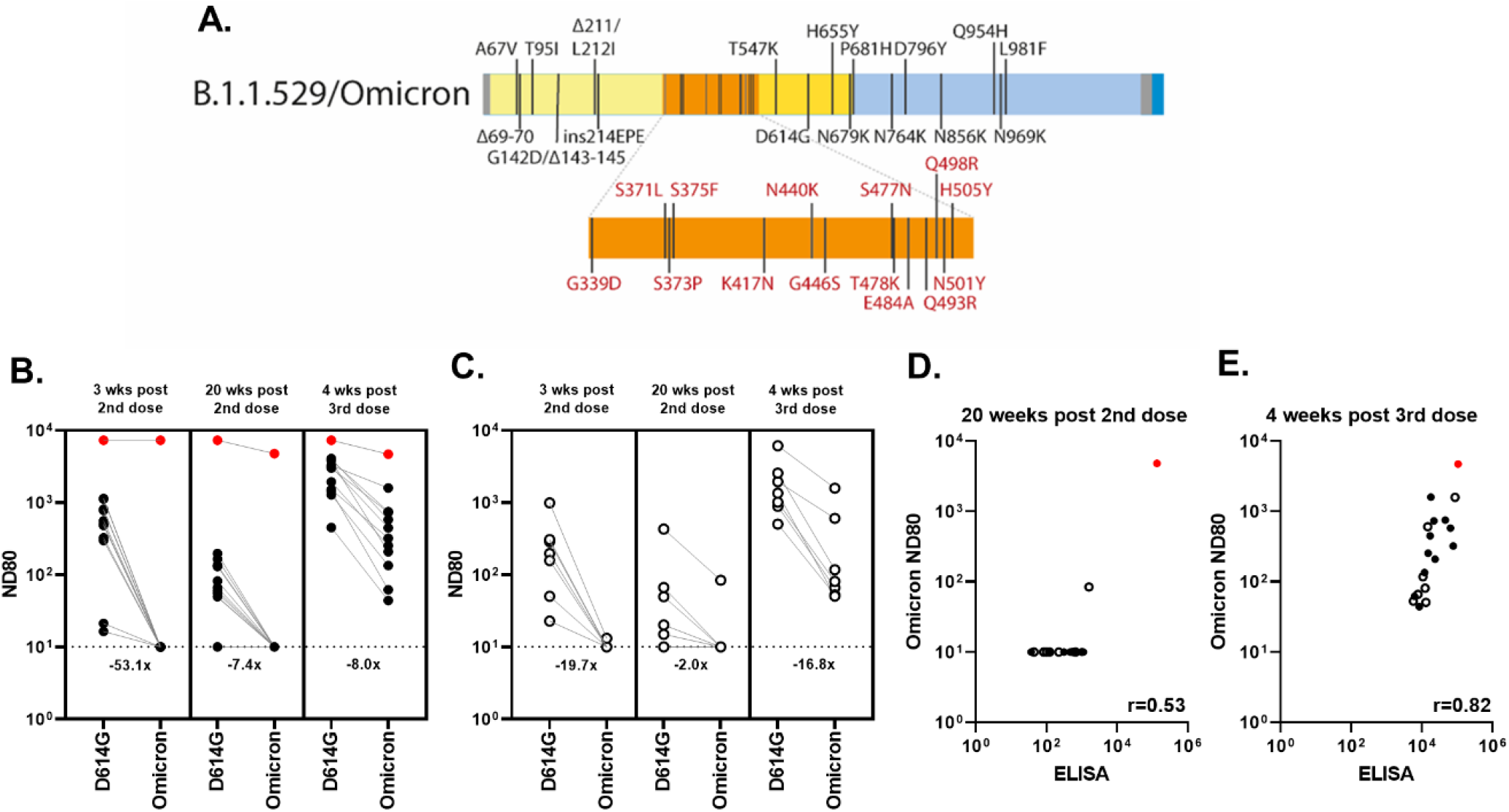
Neutralising antibody responses to Omicron after a 3rd booster dose of BNT162b2. **(A)** Schematic illustrating the spike mutation profiles of BA.1 (Omicron). Neutralisation titres calculated using pseudotypes bearing the SARS-CoV-2 D614G or Omicron spike and sera from a cohort of BNT162b2 vaccinated individuals (n=19), recruited as part of the UK CONSENSUS trial, aged 70-79 (n=12, solid circle symbols) **(B)** or 80-89 (n=7, open circle symbols) **(C)**. Sera samples were taken from the same volunteers at the indicated times post 2nd or 3rd doses of vaccination. Symbols in red represent samples taken from individuals who tested positive for SARS-CoV-2 Nucleoprotein by ELISA, indicative of previous infection. Titres are expressed as serum fold-dilution required to achieve 80% virus neutralisation, with the titre (ND80) calculated by *xy* interpolation from the mVNT data series (dilution, *x* versus luciferase activity, RLU, *y*). Fold changes in median ND80 between D614G and Omicron are indicated. The detection limit of the assay is indicated with a dotted line. The correlation between Omicron ND80 and S ELISA (Roche S) titres recorded from each volunteer, was examined at 20 weeks post 2nd dose **(D)** and 4 weeks post 3rd dose (both n=19) **(E)**, with statistical analysis of the matrix performed using a nonparametric Spearman correlation (r).

## Discussion

At a global level many elderly populations have been protected from COVID-19 by the implementation of mass vaccination. However, lower overall Ab responses and concerns of waning immunity in elderly vaccinees have highlighted the potential impact of antigenically distinct variants on controlling this disease. Using sera from an elderly cohort double-vaccinated with BNT162b2 we demonstrated significantly reduced neutralisation of the Delta and Beta VOCs, which is compounded by waning immunity (Figures 1-2); data which correlates with previously published findings. Noori et al., performed a systematic review of the potency of BNT162b2 vaccine-induced nAbs against SARS-CoV-2 VOCs from 36 different publications and identified a similar trend, with Beta being the least sensitive to neutralisation and Delta having intermediate sensitivity [22]. Similarly, a Delta-focused review by Bian et al., examining a broader selection of studies on BNT162b2 or ChAdOx1 vaccinees, as well as convalescent individuals, identified the same pattern of VOC neutralisation [23]. Indeed, across the collated studies there were numerous examples of Beta being neutralised >10-fold less efficiently than D614 or D614G (22.11-fold [24], 11.13-fold [25] and 13.32-fold [26]). Focusing on the study by Liu et al. [26], they identified a 7.56-fold reduction in neutralisation of Beta in BNT162b2-vaccinated individuals when compared to a virus isolated early in the pandemic. Differences in fold-reduction can be attributable to experimental variation (lab-to-lab, live virus vs. pseudotype) or specific characteristics of the cohort, such as age. For example, the BNT162b2-vaccinated cohort analysed by Liu et al. represents health care workers with a mean age of 37 (range 22-66), whereas our study targeted an elderly cohort aged 70-89. One interpretation is that the low nAb titres we and others have seen in elderly vaccinees [27, 28] may contribute disproportionately to the lack of neutralisation of antigenically distinct SARS-CoV-2 variants. Confirming this hypothesis would require detailed comparison of the polyclonal B-cell response in various age-groups; however, Greaney et al., demonstrated the response is skewed to a single class of antibodies which target an epitope in the RBD encompassing position 484, which is substituted in Beta (E484K) [29]. One limitation of our study is that for some VOCs, e.g. Beta, almost all the sera dropped below the limit of detection of our assay (ND80 of 10), providing less granularity, complicating comparisons with other studies, and obfuscating correlations with more sensitive Ab-detection assays such as the RBD-ELISA.

Elsewhere, our data on Omicron neutralisation following 2 doses of BNT162b2 fits with recently reported findings. Dejnirattisai et al., Cameroni et al., and Garcia-Beltran et al. all showed drops in neutralisation of >30-fold [18-20, 30]. However, it is clear from our data, and that of Garcia-Beltran et al., [20] that boosting with a 3^rd^ dose of BNT162b2 generates a much higher overall titre of neutralising antibodies to D614G (median ND80 ≥3070.9 in the 70-79 age group, Figure 4B) and that these high titres enhance cross-protection against Omicron and mitigate against the significant drops in neutralisation seen after two doses, highlighting the important role of boosters in providing robust long-term immunity to SARS-CoV-2.

A smaller pool of antigenicity data is available on the wide range of other variants (VUIs/VUMs) that have emerged since the beginning of the pandemic. Our data highlighted several important antigenically-divergent variants, in particular C.1.2 and B.1.638 (Note: B.1.638 is from a small isolated outbreak; n=13), both initially detected in South Africa [31]; B.1.621 (Mu), first detected in Colombia [32] and P.3 (Theta) – isolated in the Philippines [33]. We identified a 3.0-fold and 10.8-fold reduction in neutralisation for Mu and C.1.2, respectively (Figure 3B), and antigenic distances of 4AU (C1.2) and 3.3AU (Mu) (Figure 3C), which are concordant with observations by Tada et al. in BNT162b2-vaccinated individuals (Mu, 6.8-fold reduction; C.1.2, 7.3-fold) [34]. The same authors showed similar Mu and C.1.2-specific reductions in neutralisation in sera from convalescent individuals as well as mRNA-1273 (Moderna) vaccinees [34]. There was less agreement with the study by Uriu et al; these authors showed that Mu was more antigenically diverse than Beta (9.1-fold vs. 7.6-fold reduction with BNT162b2-vaccine serum) [35]. Similarly, Arora et al. identified that A.30 was more antigenically distinct from Beta when tested with BNT162b2 sera (4.6-fold vs. 3.3-fold) [36], whilst in our study A.30 resulted in only a 2.0-fold drop in neutralisation and a position only 2.5AU from D614G (Figure 3B). As discussed above these differences might be attributable to laboratory or cohort-specific factors. Nevertheless, the main strength of our approach has been to use a single set of sera to compare the relative neutralisation of VOCs and VUIs/VUMs (Figure 3).

As summarised by Harvey et al., [37] our molecular understanding of SARS-CoV-2 variant spike mutations and immune escape is now relatively well-established, permitting the mechanistic contextualisation of these neutralisation data. The most pertinent question raised by our data set is, ‘why are Omicron and Beta so bad?’. From a variant monitoring perspective, the answers to this question can be used as a framework to understand other SARS-CoV-2 variants, old or new. Since Omicron has so many changes in its spike protein, it is difficult to draw any meaningful conclusions without doing targeted mutagenesis. However, a number of the other variants share enough similarity to Beta that useful comparisons and conclusions can be drawn by correlating individual mutations to varied antigenicity. Compared to variants such as Omicron or B.1.638, the spike mutation profile of Beta is relatively simple, with only 9 changes (Figure 3A). Focusing on the RBD, the amino acid changes K417N and E484K are well associated with antigenic escape from monoclonal antibodies and the polyclonal B-cell response [22, 23, 29]. Changes at position 417 affect class 1 Ab binding while 484 modifications affect the epitope bound by class 2 Abs, which dominate the polyclonal response to the RBD [37]. The importance of these particular RBD-changes to Ab-binding was confirmed by our RBD-ELISA (Fig.2E-F and Fig.3D). Of the single amino acid mutated RBDs we assessed, only those containing K417N (1.2 fold) or E484K/D (E/K, 1.6-fold; E/D, 1.4-fold) significantly reduced Ab-binding, when compared to WT. The E484D change, although not present in any variant we analysed, has previously been implicated in escape from monoclonal nAbs [37]. Combining these changes with N501Y in the Beta RBD appeared to be additive and led to a concomitant 2.2-fold reduction in Ab-binding, indicating that the loss of Beta VOC neutralisation we, and others [22], have observed (Figure 2B-C) is due to a reduction in class 1 and 2 nAb binding. Interestingly, the magnitude of reduction for the complete Delta RBD (1.1-fold) or Delta-specific single amino acid RBD substitutions was lower (L452R, 1.1-fold) or the same as K417N (T478K, 1.2-fold) and non-significant in our studies (Figure 3D). This might explain the more moderate reductions in neutralisation seen with this VOC (Figure 2B-C) or point to another region of the Delta spike being responsible for antigenic variance. Further, our RBD-ELISA data on E484Q, which only moderately reduced Ab-binding (1.1-fold) might explain the relatively minor effects of B.1.617.3 on neutralisation (1.6-fold reduction, relative to D614G, Figure 3B).

Another interesting observation was to compare the contribution of 484 changes to neutralisation when in the context of other RBD changes. When present as the only RBD mutation (AT.1, 1.7-fold and 2.2AU; B.1.1.318, 2.9-fold and 3 AU), or with additional RBD changes which we demonstrated do not significantly affect Ab-binding in RBD-ELISAs, e.g. L452R (B.1.617.3, 1.6-fold and 1.9AU), E484K had only moderate effects on pseudotype neutralisation. However, the addition of N501Y (which is also present in Beta, Mu, Theta and C.1.2) seemed to enhance escape from neutralisation, e.g., P.3/Theta (7.2-fold, 3.5AU). Although there is some evidence that N501Y modifies antigenicity [38], the presence of this mutation in Alpha does not appear to affect its neutralisation (Figure 2B-C, [39]). Moreover, this substitution has most frequently been mechanistically linked to an increase in affinity for ACE2 [40]. In the context of neutralisation of E484-mutated variants it may be that the increased affinity of N501Y for ACE2 has a compound effect on class 2 antibodies trying to bind spike, as these antibodies’ affinities for spike have already been reduced by, for example, the E484K substitution. We recently showed that the RaTG13 spike is efficiently neutralised by SARS-CoV-2-specific sera, which was surprising given the large number of RBD substitutions between these two spike proteins [41]. However, the affinity of RaTG13 spike for human ACE2 is markedly lower than SARS-CoV-2 spike [42] - it is likely therefore that a limited pool of higher affinity antibodies to RaTG13 spike can effectively outcompete this interaction, permitting neutralisation, with perhaps the opposite being true in a N501Y-containing SARS-CoV-2 spike. Making conclusions on Beta mutations which lie outside of the RBD is slightly more challenging, with C1.2 or Mu/B.1.621 being perhaps the best comparator. The Mu + K417N variant spike we analysed (3.2-fold reduction in neutralisation and 2.1 AU from D614G) has almost the same RBD as Beta (albeit Mu additionally has R346K); however, the N-terminal domains (NTD) are distinct. Beta has a deletion between amino acid positions 242-244 [43], recurrently deleted region 4 (RDR4), which corresponds to exposed loops on the surface of the spike NTD. RDR4 deletions have previously been associated with the loss of monoclonal antibody binding (4A8)[43]. Likewise, L18F also sits within the NTD supersite and its modification may also affect antibody binding.

To summarise, the significant antigenic escape properties of Beta and Omicron are likely the result of a combination of changes to the spike protein, which act in synergy and in an additive manner to avoid Ab-recognition, with the other variants we tested lacking some or all of these features. These hypotheses could be tested by introduction of K417N into C.1.2, the 242-244 deletion into Mu + K417N, or by selected deletion of mutations in Omicron. Of note, this research should only be performed in the context of spike expression plasmids for pseudotyping, and not in gain-of-function experiments with recombinant viruses. Of note, it is also worth highlighting that antigenic changes to spike are not the only defining feature of SARS-CoV-2 variants. For instance, Alpha and Delta have risen to global dominance over Beta, likely because of changes elsewhere in spike that affect infectivity and transmissibility [44] or indeed changes elsewhere in the genome, which affect other phenotypic characteristics of the virus such as innate immune regulation [45]. Nevertheless, these detailed antigenic data can be used as a reference for risk assessment of emerging SARS-CoV-2 variants.

Focusing on the UK situation momentarily: the UK has extensive monitoring programmes for SARS-CoV-2 which are combined with genomic sequencing to identify novel variants as and when they emerge or are introduced [13]. From the summer of 2021 onwards a sub-lineage of Delta, AY.4.2, has increased in prevalence, and in the last 28 days (up to 10-12-2021) has accounted for 18.2% of the genotyped isolates (https://sars2.cvr.gla.ac.uk/cog-uk/). This variant, which early data indicates may have higher transmissibility [46], has two additional changes in the spike NTD, Y145H and A222V. Interestingly, despite these additional changes, pseudotypes bearing the AY.4.2 spike as well as Delta + A222V alone were more efficiently neutralised by sera from our BNT162b2-cohort (Figure 3) when compared to Delta (albeit not significantly; Wilcoxon matched-pairs signed rank test). These data are similar to those reported by Lassauniere et al., who also examined BNT162b2 mRNA vaccine-elicited sera [47]. A222V is the second most common substitution in SARS-CoV-2 Spike (after D614G), yet to date had no established phenotypic impact on spike function [37]. It remains unclear why A222V increases the neutralisation of Delta; however, this could relate to increased accessibility of the RBD to nAbs. It remains to be seen how the prevalence of AY.4.2 will be affected by the introduction of Omicron to the UK; however, epidemiologists are predicting that Delta and its sub-lineages will soon be replaced by this new variant [13].

Another pertinent question that our data set raises is whether spike-based ELISA titres correlate well with virus (or pseudovirus) neutralisation titres. From a practical perspective, if ELISAs can be used to establish an immune threshold that in turn reliably correlates with a certain level of nAbs, then that would be advantageous for clinical management of COVID-19. This is especially true if a threshold can easily be established for each variant as and when they emerge. Unfortunately, whilst our data on a BNT162b2-vaccinated elderly cohort identified a reliable correlation between WT (D614 or D614G), Alpha and Delta, it was disrupted by more antigenically distinct variants like Beta (Figure 2) and Omicron (Figure 4D). Whilst the RBD-ELISA with a Beta RBD was useful for mechanistically underpinning our understanding of ‘why Beta is so bad’, and the antigenic relationships were largely consistent between mVNT and ELISA using antigenic maps, the results did not improve the correlation with neutralisation titres, although a better correlation exists with higher titre sera. However, a major limitation of our findings is that our mVNT was clearly less sensitive than the ELISA or RBD-ELISA, with many Beta mVNTs falling below the limit of detection (<10 ND80). A younger cohort, one sampled from vaccinees who had the extended interval between 1^st^ and 2^nd^ dose [48], or people who receive a 3^rd^ dose (Figure 4) who collectively have higher overall titres, might represent a better benchmark for understanding and testing this correlation in more detail.

A younger cohort, including recipients of other vaccines, could also improve the resolution of the antigenic maps. Antigenic cartography offers great potential as a robust and repeatable way to measure antigenic changes among variants, and test hypotheses on the effect of individual and combinations of spike protein mutations [49, 50]. However, using sera with similar neutralisation profiles and from volunteers with unknown history of exposure to other viruses limits the resolution of the maps [51]. Using sera from other species with a known exposure history has also demonstrated its utility in rigorous comparison of antigenic distances, overcoming these issues [52].

## Concluding remarks

In summary, our data highlight the propensity of certain SARS-CoV-2 variants to partially avoid vaccine-derived immunity. The BNT162b2-vaccinated elderly cohort we investigated showed evidence of waning immunity at 20-weeks post-vaccination, which could potentially exacerbate this escape. However, high titres could be rescued with a 3^rd^ dose, which provided cross-protective immunity against Omicron. Developing a better understanding of how these titres relate to a well-defined correlate of immunity will be an important step in understanding the wider implications of this data on the management of COVID-19 and whether the risk of breakthrough infections, hospitalisation and deaths is increased, either by waning immunity or new variants. Interestingly, three of the vaccinated volunteers in our study were SARS-CoV-2 N Ab positive (Elecsys® Anti-SARS-CoV-2 N, Roche), indicative of previously resolved infections. The nAb and ELISA titres from two of these individuals were the highest in our cohort, supporting observations that infection and vaccination provide a robust Ab response to SARS-CoV-2 [53]. Clearly, we can achieve similar titres through boosting (Figure 4); however, whether boosting provides as broad a response as natural infection in this context remains to be determined. Interestingly, it is clear from various studies that responses to infection and/or vaccination can be variant-specific, and that neutralisation of homologous variants is superior to heterologous [54, 55]. For example, Liu et al., showed that sera from convalescent individuals who had been infected with Beta and Gamma VOCs showed superior responses to these matched variants in VNTs, but reduced neutralisation of Delta [26]. For Delta, which does not appear that antigenically distinct from the ‘vaccine strain’, it may not be necessary to tailor the Ab response with variant-specific boosters, a conclusion that our data indicates might also be true for Omicron. To conclude, research comparing the antigenicity of SARS-CoV-2 variants and elucidating the role of individual mutations is vital in the management of the Covid-19 pandemic and in predicting the outcome of new variants. At a clinical level this information can be leveraged by policymakers to determine the most appropriate vaccination strategy to protect the most vulnerable groups in society.

## Data Availability

All data produced in the present study are available upon reasonable request to the authors.

## Acknowledgements

We would like to acknowledge the whole of the UKHSA (formerly PHE) CONSENSUS team, as well as the vaccinated volunteers for their help and participation in supporting this study. This work was supported by the MRC funded G2P-UK National Virology Consortium; G2P-UK; A National Virology Consortium to address phenotypic consequences of SARS-CoV-2 genomic variation (MR/W005611/1). JN, NT, DB, AE, BC and DB were also funded by The Pirbright Institute’s BBSRC institute strategic programme grant (BBS/E/I/COV07001, BBS/E/I/00007031, BBS/E/I/00007038, BBS/E/I/00007039 and BBS/E/I/00007034) with NT receiving studentship support from BB/T008784/1. CB and DH were supported by funding from the European Union’s Horizon 2020 Research and Innovation programme under grant agreement No 773830: One Health European Joint Programme. We would also like to acknowledge the National Institute for Communicable Diseases (NICD) and the KZN Research Innovation and Sequencing Platform (KRISP), as part of the Network for Genomic Surveillance in South Africa, for depositing the B.1.638 sequences, and the NICD, in particular Daniel G. Amoako and Josie Everatt for providing feedback on the manuscript. The Sequencing activities at the NICD were supported by: a conditional grant from the South African National Department of Health as part of the emergency COVID-19 response; a cooperative agreement between the National Institute for Communicable Diseases of the National Health Laboratory Service and the UK Department of Health and Social Care, managed by the Fleming Fund and performed under the auspices of the SEQAFRICA project.

## Methods

### Participants and Ethical Statement

Healthy participants were recruited as part of the “COVID-19 vaccine responses after extended immunisation schedules” (CONSENSUS) study [21]. Participants aged 70-90 years in January 2021 were recruited through North London primary care networks. Sera samples used in this study were taken at 3 and 20 weeks after 2 doses of Pfizer/BioNTech BNT162b2 (Mainz, Germany) COVID-19 vaccine given at a 3-week interval, as well as 4 weeks post 3^rd^ dose. The protocol was approved by Public Health England Research Ethics Governance Group (reference NR0253; 18/01/21). All sera were heat inactivated at 56 degrees for 2 hours prior to use.

### Cells

Human embryonic kidney (HEK) 293T cells were used to generate lentiviral pseudoparticles bearing the SARS-CoV-2 spike. HEK293T cells stably expressing human angiotensin converting enzyme 2 (hACE2) under 1µg/mL puromycin (Gibco) selection were used for neutralisation assays [56]. Cells were maintained in Dulbecco’s Modified Eagle Medium (Sigma-Aldrich) supplemented with 10% FBS (Life Science Production, UK), 1% 100 mM sodium pyruvate (Sigma-Aldrich), 1% 200 mM L-glutamine (Sigma-Aldrich), and 1% penicillin/streptomycin, 10,000 U/mL (Life Technologies) at 37°C in a humidified atmosphere of 5% CO2.

### Plasmids

Mutant SARS-CoV-2 expression plasmids were generated by site-directed mutagenesis, by using the QuikChange Lightning Multi Site-Directed Mutagenesis Kit (Agilent) or were synthesised by Geneart (Thermo Fisher). All SARS-CoV-2 spike expression plasmids were based on a codon optimised Wuhan-Hu-1 reference sequence (Genbank ID NC_045512.2) [57], with the additional substitutions K1255*STOP (also referred to as Δ19 mutation or cytoplasmic tail truncation). A list of the SARS-CoV-2 spike variants and their associated mutations can be found in Supplemental Table 1. Some substitutions here differ from the lineage defining sequence for the named variant (Pango); these were included as they were substitutions that were highly sampled in submitted sequences and predicted to be a plausible worst case antigenic escape for that lineage.

### Generating lentiviral based pseudotypes bearing the SARS-CoV-2 spike

Lentiviral-based SARS-CoV-2 pseudotyped viruses were generated in HEK293T cells incubated at 37°C, 5% CO_2_ as previously described [58]. Briefly, cells were transfected with 900 ng of SARS-CoV-2 spike (see Supplemental Table 1), 600 ng p8.91 (encoding for HIV-1 gag-pol), 600 ng CSFLW (lentivirus backbone expressing a firefly luciferase reporter gene) with PEI (1 µg/mL) transfection reagent. Supernatants containing pseudotyped SARS-CoV-2 were harvested and pooled at 48 and 72 hours post transfection, centrifuged at 1,300 x *g* for 10 minutes at 4 °C to remove cellular debris and stored at -80 °C. SARS-CoV-2 pseudoparticles were titrated on HEK293T cells stably expressing human ACE2 S-CoV-2 pps and infectivity assessed by measuring luciferase luminescence after the addition of Bright-Glo luciferase reagent (Promega) and read on a GloMax Multi+ Detection System (Promega).

### Micro neutralisation test (mVNT) using SARS-CoV-2 pseudoparticles

Sera was diluted 1:5 in serum-free media in a 96-well plate in triplicate and titrated 3-fold. A fixed titred volume of SARS-CoV-2 pseudoparticles was added at a dilution equivalent to 10^5^-10^6^ signal luciferase units in 50 µL DMEM-10% and incubated with sera for 1 hour at 37 °C, 5% CO_2_ (giving a final sera dilution of 1:10). Target cells expressing human ACE2 were then added at a density of 2 *x* 10^4^ in 100 µL and incubated at 37 °C, 5% CO_2_ for 48 hours. Firefly luciferase activity was then measured after the addition of BrightGlo luciferase reagent on a Glomax-Multi^+^ Detection System (Promega, Southampton, UK). Pseudotyped virus neutralisation titres were calculating by interpolating the point at which there was 80% reduction in reduction in luciferase activity, relative to untreated controls, neutralisation dose 80% (ND_80_).

### ELISA RBD

Antibody to the RBD of SARS-CoV-2 spike protein was measured using an in-house indirect IgG RBD assay as described previously [59, 60]. Briefly, commercially synthesised recombinant RBD subunit spike(Arg319-Phe541(V367F); SinoBiological Inc, Hong Kong, PRC) with a C-terminal mouse Fc tag, was coated onto 96-well microtiter plates at 20 ng per well at 4°C–8°C for a minimum of 16 hours. After washing and blocking, sera were analysed at a dilution factor of 1 in 100 by serially diluting each serum sample starting at 1:100, (6-fold with the highest dilution achieved 129600). The binding of IgG on the plate surface was detected by using an anti-human IgG horseradish peroxidase antibody conjugate (Sigma-Aldrich, Poole, UK) and 3,3′,5,5′-Tetramethylbenzidine (Europa Bioproducts Ltd, Ely, UK). We analysed samples in duplicate and evaluated optical density at 450 nm (OD_450_) Samples were analysed in the presence of known positive controls (collected from individuals with confirmed SARS-CoV2 infection) and a calibrator sample (“negative” added to four wells; collected prior to the pandemic). Titres are expressed as serum fold-dilution required to achieve a T/N (test OD to negative OD) of 5 (T/N = 5 serves as cut-off for positive samples) by xy interpolation from the RBD data series (dilution, x versus OD450, y). Samples which were below the cut-off in the initial dilution (ie negative), were expressed as <100.

### ELISA N and S Roche

Sera samples were tested by commercial ELISA as previously described [21]. Nucleoprotein (N) antibodies were measured as a marker of previous SARS-CoV-2 infection (Anti-SARS-CoV-2 total antibody assay, Roche Diagnostics, Basel, Switzerland) and spike (S) protein antibodies were measured as an indication of infection or vaccination (Elecsys Anti-SARS-CoV-2 S total antibody assay, Roche Diagnostics: positive ≥ 0.8 arbitrary units (au)/mL to assess vaccine response).

### Infectious SARS-CoV-2 VNT

Sera were serially diluted 1:2 in media containing 1% foetal calf serum (FCS) and incubated with 112.5 plaque forming units (PFU) of SARS CoV-2 (hCoV-19/England/02/2020, EPI_ISL_407073, or Beta (B.1.351), kindly provided by Public Health England) for one hour at 37 °C, 5% CO_2_. 75 PFU of neutralisation mixture in a total of 200µl were then added to 96-well plates containing a ∼80-90% confluent monolayer of Vero-E6-TMPRSS2 (a gift from Stuart Neil, KCL, London) cells were incubated for 6 days at 37 °C, 5% CO_2_ in quadruplicate per serum sample. Inoculum was then removed, and cells were fixed with formalin for 30 minutes before staining with 0.1% toluidine blue in PBS. ND_80_ titres were calculated using a Spearman and Karber formula. Thawed virus aliquots used for VNT were back-titrated 1:10 by TCID_50_ on VERO-E6-TMPRSS2 cells to confirm the titre at time of use.

### Cartography

Antigenic maps were made with antigenic cartography techniques described previously [50], implemented in the R package *Racmacs* (version 1.1.12; https://acorg.github.io/Racmacs/). In brief, titres were first converted into serum-antigen target distances, and sera and antigens were then positioned on a map in a way that minimised the difference (residual sum of squares) between target distances and corresponding map distances, using multidimensional scaling. The target distance for each serum-antigen pair was calculated by subtracting the logarithm (log_2_) of the titre, from the highest log_2_(titre) for the serum against any antigen; thus, higher reciprocal titres resulted in shorter target distances. Multidimensional scaling was carried out with 1000 random restart optimisations, to avoid local optima and increase the likelihood of finding the best fit to the measured titres. The resulting maps were ordered according to total error, and compared for self-consistency; the figures and descriptions in this manuscript pertain to the maps with the lowest total error. Antigenic distances were measured from the lowest error antigenic map.

The antigenic map of the ND80 titres was made based on titres from a subset of 14 sera: two sera were removed before mapping, as their titres were consistently below or only marginally above the detection limit, and they did therefore not contain valuable information for cartography. ND80 titres from different experiments were merged without normalisation procedures. For each serum, a single overall log_2_(titre) to D614G and B.1.617.2/Delta was calculated by taking the mean of the log_2_(titre) values for these antigens across experiments. Such average log_2_(titre) values were excluded (replaced with NA/’missing value’) in case the standard deviation of log_2_(titre) values for the antigen was equal to or exceeded 1.

To determine the optimal number of dimensions for representing the data, prediction experiments were performed: antigenic maps were made while omitting a random 10% of titres; the excluded titres were predicted according to their relative positions in the map; and the predicted titres were then compared to the actual titres (on a log scale). Antigenic maps were made in two, three, four and five dimensions, using 1000 optimizations; for each dimension, 100 prediction tests were performed. The mean root mean square error (RMSE) associated with the prediction of ND80 titres was 0.94 (SD: 0.15) for detectable titres and 1.32 (SD: 0.68) for non-detectable titres (i.e., below the limit of detection, <10), for two dimensions; 0.97 (SD: 0.14) and 1.37 (SD: 0.61) for three dimensions; 0.96 (SD: 0.14) and 1.34 (SD: 0.60) for four dimensions; and 0.95 (SD: 0.14) and 1.35 (SD: 0.58) for five dimensions. The mean RMSE associated with the prediction of RBD-ELISA titres was 0.55 (SD: 0.16) for detectable titres and 1.27 (SD: 0.13) for non-detectable titres (i.e., below the limit of detection, <100), for two dimensions; 0.56 (SD: 0.15) and 1.26 (SD: 0.10) for three dimensions; 0.56 (SD: 0.14) and 1.25 (SD: 0.10) for four dimensions; and 0.56 (SD: 0.15) and 1.25 (SD: 0.11) for five dimensions. Overall, for each dataset, the mean RMSE was similar across dimensions, and in each case the mean RMSE for prediction of detectable titres corresponded to less than a twofold dilution (one antigenic unit). Therefore, we considered two-dimensional maps sufficient for representing the SARS-CoV-2 antigenic data.

