## Supplemental Figures 1-5 for "Neutralising antibody activity against SARS-CoV-2 variants, including Omicron, in an elderly cohort vaccinated with BNT162b2"

### A. Dilution series: 1:40 – 1:2560

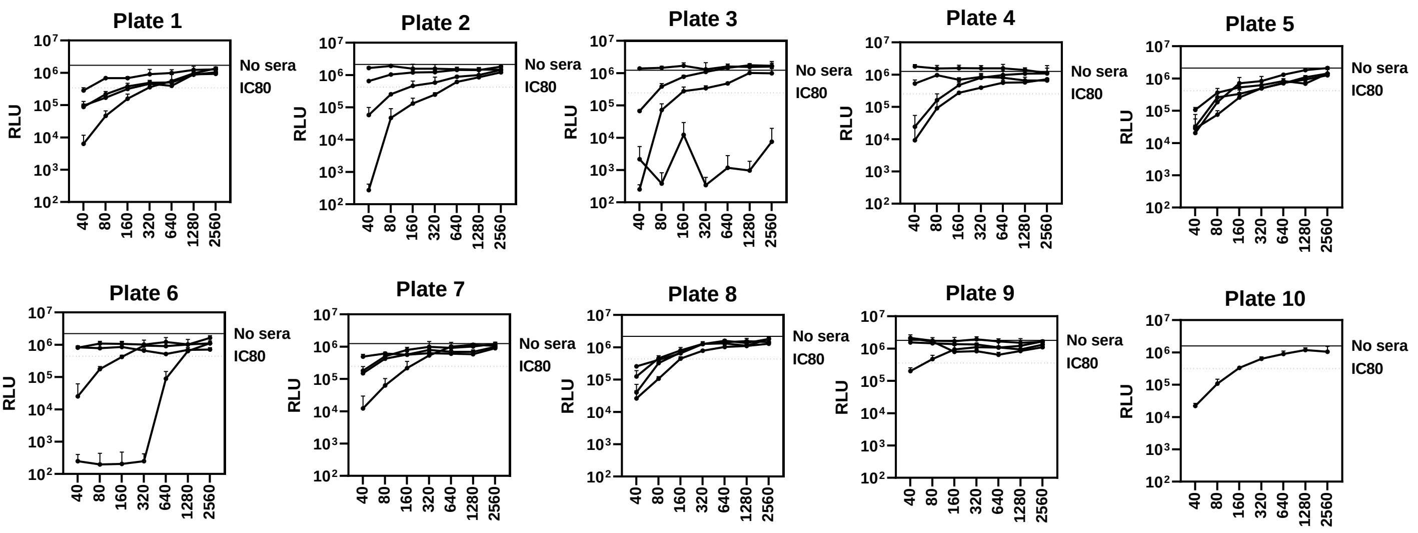


### B. Dilution series: 1:10 – 1:80

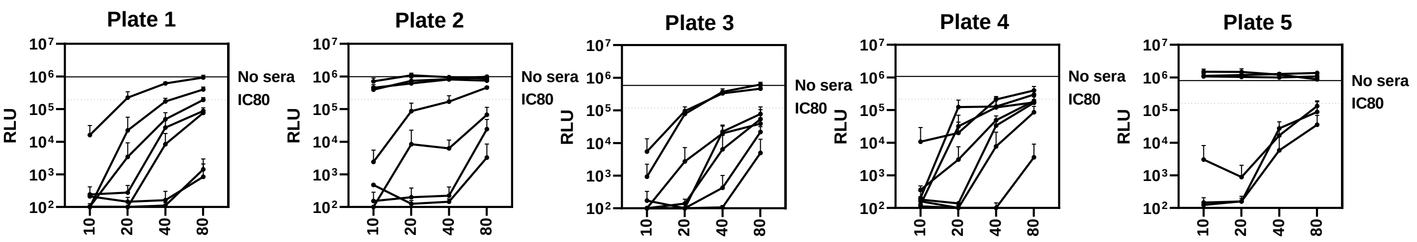


C.

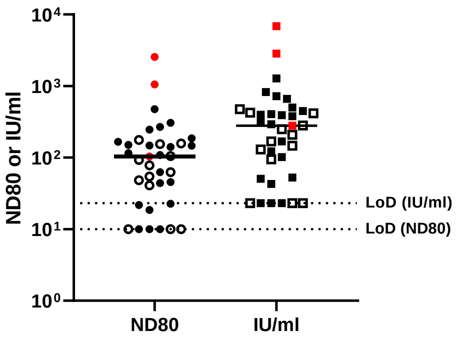


**Supplemental Figure 1: Micro virus neutralisation test (mVNT) data examining the neutralising antibody responses generated following BNT162b2 vaccination.** Neutralisation titres in **Fig.1-3** were calculated using pseudotypes bearing various SARS-CoV-2 Spikes and sera from a cohort of BNT162b2 vaccinated individuals (n=37), recruited as part of the UK CONSENSUS trial, aged 70-89 (n=37). Titres are expressed as serum fold-dilution required to achieve 80% virus neutralization, with the titre (ND80) calculated by *xy* interpolation from the mVNT data series (dilution, *x* versus luciferase activity, RLU, *y*). Provided in **(A)** for illustrative and representative purposes are the graphed data from mVNTs (n=37) of WT D614 (Wuhan) pseudotypes using the sera collected from this cohort at 3 weeks post 2$^{nd}$ dose, diluted 1:40 to 1:2560 (**Fig.1A-B**). **(B)** Results from repeated mVNTs with the same sera panel, using instead a dilution series of 1:10 to 1:80 to detect lower levels of neutralizing antibodies. Each graph represents data from a separate 96-well plate used for mVNT, with the plate specific 'no sera' and 'IC80' values highlighted. **(C)** mVNTs for these samples were run at the same time as the WHO's International Standard (IS) for SARS-CoV-2 serological assays (NIBSC code: 20/136), to calculate IU/ml values across the experiment. Both the original ND80 titre (round symbol) and IU/ml (square symbol) values are plotted (ages 70-79, n=24, solid symbols; 80-89, n=13, open symbols). Symbols in red represents samples taken from individuals who tested positive for SARS-CoV-2 Nucleoprotein by ELISA, indicative of previous infection. The relative detection limit of each assay is indicated.

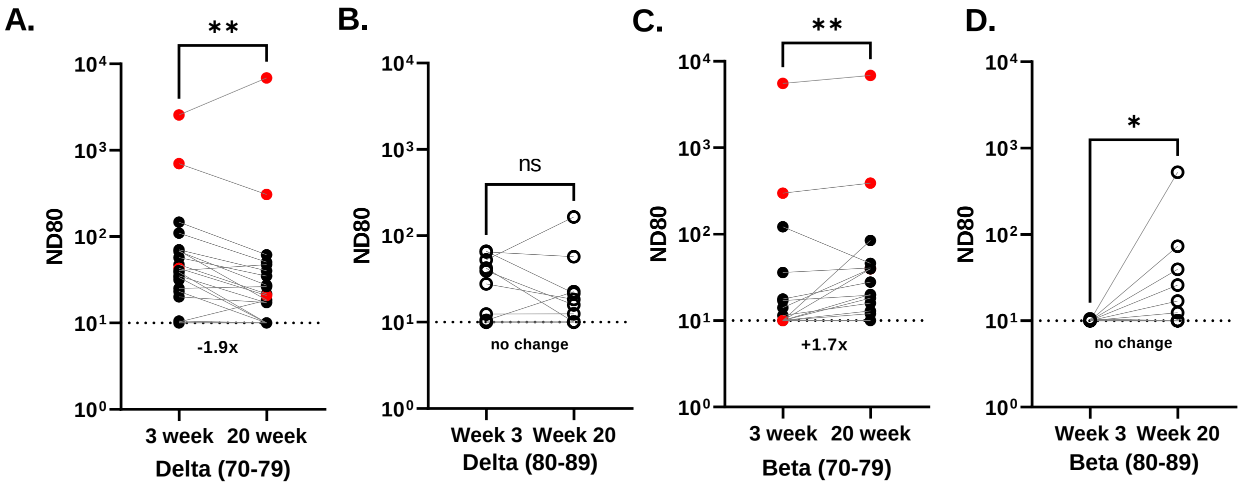


**Supplemental Figure 2: Comparison of neutralising antibody titres against SARS-CoV-2 VOCs at 3- and 20-weeks post 2nd dose of BNT162b2.** Neutralisation of pseudotypes bearing the SARS-CoV-2 Delta or Beta Spike were compared in the two age-stratified cohorts, 70-79 (solid circles) (**A/C**) and 80-89 (open circles) **(B/D)**. Sera for mVNTs was collected from the same individuals at 3- (n=37 total) and 20-weeks (n=35 total) post 2nd dose. Symbols in red represents samples taken from individuals who tested positive for SARS-CoV-2 Nucleoprotein by ELISA, indicative of previous infection. Titres are expressed as serum fold-dilution required to achieve 80% virus neutralization, with the titre (ND80) calculated by *xy* interpolation from the mVNT data series (dilution, *x* versus luciferase activity, RLU, *y*). The detection limit of the assay is indicated with a dotted line. Statistical comparison of ND80 titres at 3 and 20 weeks was performed using a Wilcoxon matched-pairs signed rank test (*; <0.05; **, <0.01). Fold changes in median ND80 between 3 and 20 weeks are indicated.

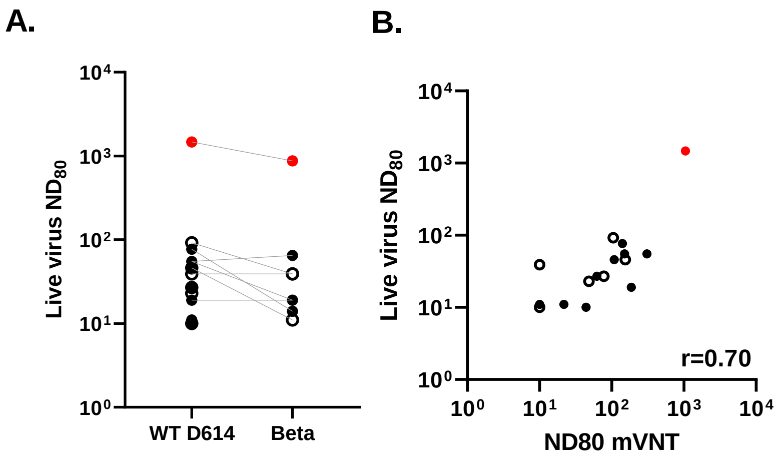


**Supplemental Figure 3: Comparison of neutralising antibody titres using pseudotype and live SARS-CoV-2 VOCs. (A)** ND80s were calculated using sera from a sub-section (n=16) of the BNT162b2-vaccinated cohort (ages 70-79, solid circles; 80-89, open circles) and live SARS-CoV-2 virus isolates (WT/ D614 or Beta). **(B)** The corresponding ND80s calculated using pseudotypes (Fig.2) were compared to these live virus ND80s, with statistical analysis of the matrix performed using a nonparametric Spearman correlation (r). Symbols in red represents samples taken from individuals who tested positive for SARS-CoV-2 Nucleoprotein by ELISA, indicative of previous infection.

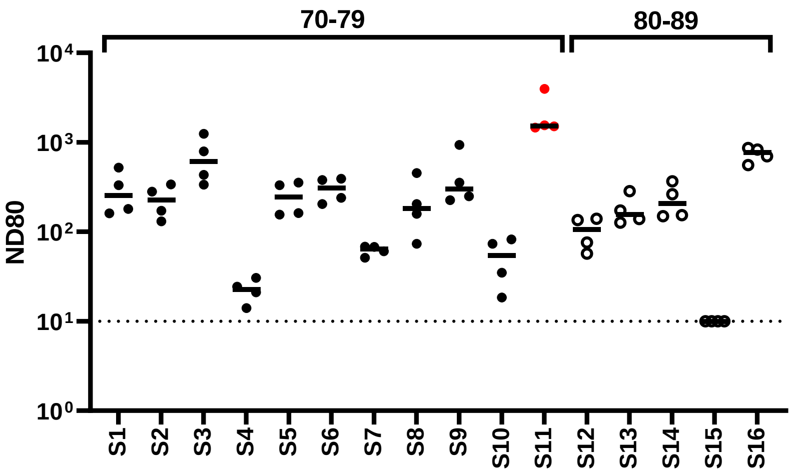


**Supplemental Figure 4: Repeat investigation of SARS-CoV-2 D614G neutralisation indicates good concordance between assays.** Neutralisation titres against D614G reported in Fig.1 and Fig.2 were repeated across four experiments with the same sub-selection (n=16) of the BNT162b2-vaccinated cohort (ages 70-79, solid circles; 80-89, open circles). Plotting of these ND80 titres indicates high concordance, and robust repeatability for the mVNT assay. Symbols in red represents samples taken from individuals who tested positive for SARS-CoV-2 Nucleoprotein by ELISA, indicative of previous infection. The detection limit of the assay is indicated with a dotted line.

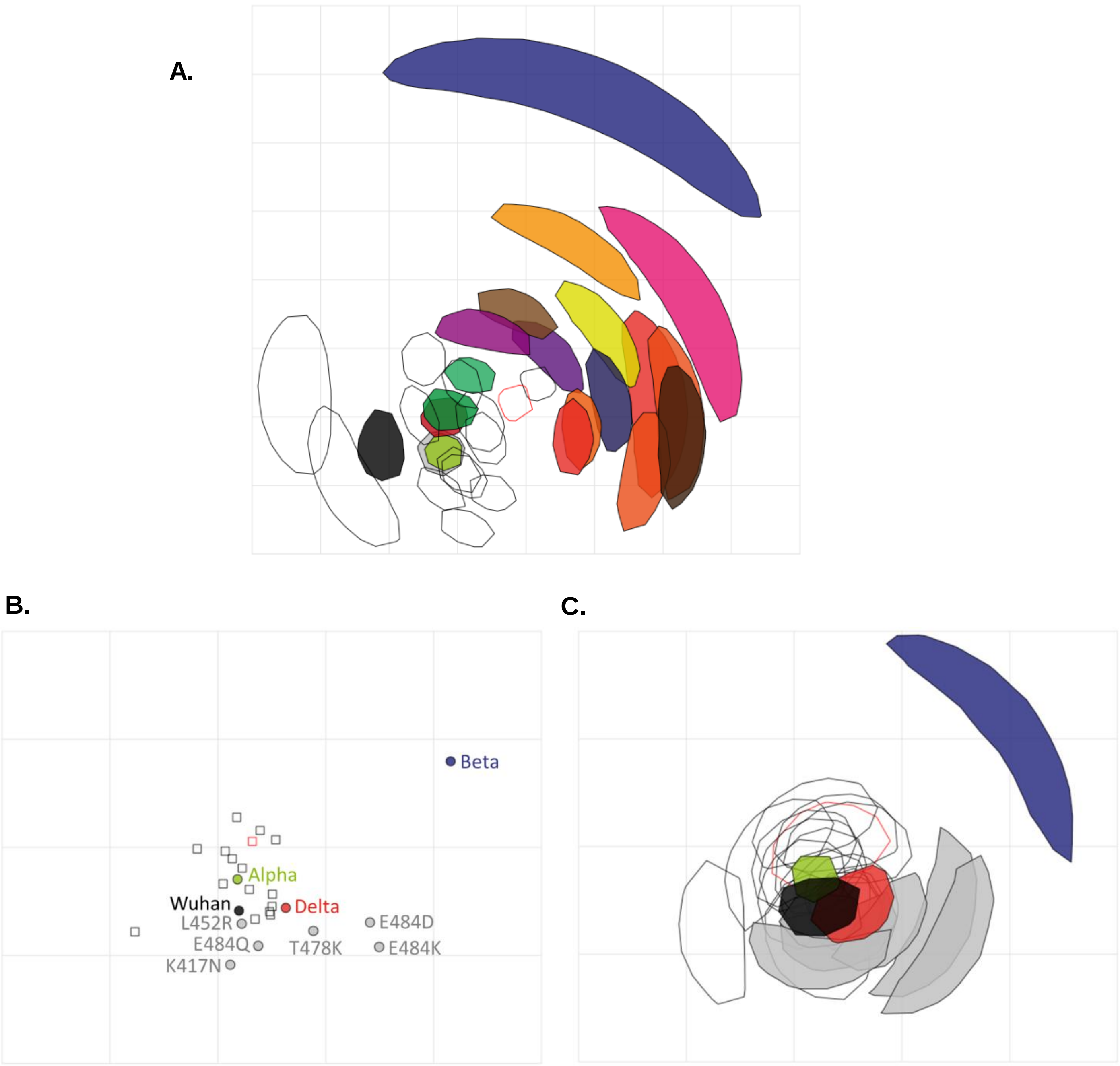


**Supplemental Figure 5: Antigenic cartography. (A)** Two-dimensional antigenic map of variants (the same map as in Figure 3C), with coordination confidence areas reflecting the uncertainty in positioning of variants and sera. Variants are represented by solid shapes, sera by open shapes (with in red a serum from an individual who tested positive for SARS-CoV-2 Nucleoprotein by ELISA, indicative of previous infection). Each shape encompasses the area on the map that the point could be located at, without increasing the error of the map by more than 1. The spacing between grid lines represents one antigenic unit, equivalent to a two-fold dilution in ND80 titres. **(B)** Two-dimensional antigenic map of variant Spike RBDs, based on the RBD-ELISA titres in Figure 3D. Multidimensional scaling was used to position the sera and Spike RBDs to best fit target distances derived from the titres. The map is the lowest error solution of 1000 optimisations. Spike RBDs are represented by solid circles (variants coloured as in Figure 3A, RBDs with individual mutations in grey), sera by open squares (with the Nucleoprotein-ELISA-positive serum in red). The spacing between grid lines represents one antigenic unit, equivalent to a two-fold dilution in RBD-ELISA titres. **(C)** The same map as (B), with coordination confidence areas reflecting the uncertainty in positioning of Spike RBDs and sera. Each shape encompasses the area on the map that the point could be located at, without increasing the error of the map by more than 1.
