## Supplemental Table 1 for "Neutralising antibody activity against SARS-CoV-2 variants, including Omicron, in an elderly cohort vaccinated with BNT162b2"

Supplemental Table 1: Spike plasmids for pseudo-typing

| Pango lineage | WHO designation | Spike mutations (compared to Wuhan-01) |
| --- | --- | --- |
| B |  | n/a |
| B.1 |  | D614G |
| B.1.1.7 | Alpha | Δ69-70, Δ144, N501Y, A570D, D614G, P681H, T716I, S982A, D1118H |
| B.1.351 | Beta | L18F, D80A, D215G, Δ242-244, K417N, E484K, N501Y, D614G, A701V |
| B.1.617.2 | Delta | T19R, G142D, Δ156-157/R158G, L452R, T478K, D614G, P681R, D950N |
| B.1.617.2 + A222V |  | T19R, G142D, Δ156-157/R158G, A222V, L452R, T478K, D614G, P681R, D950N |
| AY4.2 |  | T19R, G142D, Y145H, Δ156-157/R158G, A222V, L452R, T478K, D614G, P681R, D950N |
| B.1.214.2 |  | R214-insTDR-D215, Q414K, N450K, D614G, T716I |
| C.36.3 |  | S12F, Δ69-70, W152R, R346S, L452R, D614G, Q677H, A899S |
| C.37 | Lambda | G75V, T76I, Δ246-252/D253N, L452Q, F490S, D614G, T859N |
| B.1.1.318 |  | T95I, Δ144, E484K, D614G, P681H, D796H |
| AT.1 |  | P9L, Δ136-144, D215G, H245P, E484K, D614G, N679K/insGIAL, E780K, |
| B.1.620 |  | P26S, Δ69-70, V126A, Δ144, Δ242-244, S477N, E484K, D614G, P681H, T1027I, D1118H |
| A.30 |  | D80Y, Δ144, Δ210, D215G, Δ246-248/L249M, W258L, R346K, T478R, E484K, H655Y, P681H, Q957H |
| B.1.617.3 |  | T19R, Δ156-157/R158G, L452R, E484Q, D614G, P681R, D950N, |
| B.1.621 | Mu | T95I, Y144T/144insS/Y145N, R346K, E484K, N501Y, D614G, P681H, D950N |
| B.1.621 + K417N |  | T95I, Y144T/144insS/Y145N, R346K, K417N, E484K, N501Y, D614G, P681H, D950N |
| P.3 (B.1.1.28.3) | Theta | Δ141-143, Δ243-244, Y265C, E484K, N501Y, D614G, P681H~~,~~ E1092K, H1101Y, V1176F |
| C.1.2 |  | P9L, C136F, R190S, D215G, Δ242-243, Y449H, T478K, E484K, N501Y, D614G, H655Y, N679K, T716I |
| B.1.638 |  | L5F, Δ14-18, W64R, T95I, C136Y/Δ137, Δ141-144, W152R, Δ210, P348L, N440K, Y453F, N460K, A475V E484K, D614G, P621S, H655Y, P681H, T859N, D936G, G1219C |
| BA.1 | Omicron | A67V, Δ69/70, T95I, G142D, Δ143/145, N211I, Δ212, G339D, S371L, S373P, S373F, S477N, T478K, E484A, Q493R, G496S, Q498R, N501Y, Y505H, T547K, D614G, H655Y, N679K, P681H, N764K, D796Y, N856K, Q954H, N969K, L981F |
